# A systematic review and meta-analysis reveals long and dispersive incubation period of COVID-19

**DOI:** 10.1101/2020.06.20.20134387

**Authors:** Yongyue Wei, Liangmin Wei, Yihan Liu, Lihong Huang, Sipeng Shen, Ruyang Zhang, Jiajin Chen, Yang Zhao, Hongbing Shen, Feng Chen

## Abstract

**Background:** The incubation period of SARS-CoV-2 remains uncertain, which has important implications for estimating transmission potential, forecasting epidemic trends, and decision-making in prevention and control.

**Purpose:** To estimate the central tendency and dispersion for incubation period of COVID-19 and, in turn, assess the effect of a certain length of quarantine for close contacts in active monitoring.

**Data Sources:** PubMed, Embase, medRxiv, bioRxiv, and arXiv, searched up to April 26, 2020

**Study Selection:** COVID-19 studies that described either individual-level incubation period data or summarized statistics for central tendency and dispersion measures of incubation period were recruited.

**Data Extraction:** From each recruited study, either individual-level incubation period data or summarized statistics for central tendency and dispersion measures were extracted, as well as population characteristics including sample size, average age, and male proportion.

**Data Synthesis:** Fifty-six studies encompassing 4 095 cases were included in this meta-analysis. The estimated median incubation period for general transmissions was 5.8 days [95% confidence interval (95%CI), 5.3 to 6.2 d]. Median and dispersion were higher for SARS-CoV-2 incubation compared to other viral respiratory infections. Furthermore, about 20 in 10 000 contacts in active monitoring would develop symptoms after 14 days, or below 1 in 10 000 for young-age infections or asymptomatic transmissions.

**Limitation:** Small sample sizes for subgroups; some data were possibly used repeatedly in different studies; limited studies for outside mainland China; non-negligible intra-study heterogeneity.

**Conclusion:** The long, dispersive incubation period of SARS-CoV-2 contributes to the global spread of COVID-19. Yet, a 14-day quarantine period is sufficient to trace and identify symptomatic infections, which while could be justified according to a better understanding of the crucial parameters.

## INTRODUCTION

In December 2019, a cluster of pneumonia cases with unclear pathogenesis was reported in Wuhan, Hubei Province, China. This virus was named by World Health Organization (WHO) as the severe acute respiratory syndrome coronavirus (SARS-CoV-2) and the disease it caused was named as coronavirus disease 2019 (COVID-19) on February 11, 2020 (1). Consequently, COVID-19 was urgently classified as a Class B communicable disease and managed as a Class A communicable disease in accordance with the Law of the People’s Republic of China on the Prevention and Treatment of Infectious Disease (2). Meanwhile, the COVID-19 epidemic continued to spread around the globe, with rapid increases in case numbers in European and American countries, and a looming threat in resource-limited settings across Africa (3). The World Health Organization (WHO) declared a global pandemic on March 11, 2020. As of May 25, the pandemic had spread to 188 countries on six continents, with a total of over 5 million diagnosed cases worldwide (4).

Defining the incubation period of any infectious disease is crucial to evaluate transmission potential, estimate epidemic trends, and inform active monitoring and/or mandatory quarantine policies. The novel pathogenesis of COVID-19 has produced varied epidemiological characteristics from previous coronavirus-derived pulmonary infectious diseases, such as severe acute respiratory syndrome (SARS) and Middle East respiratory syndrome (MERS). SARS and MERS were rarely transmitted during the asymptomatic period (5, 6). In contrast, increasing evidence indicates that individuals infected with SARS-CoV-2 could be infectious during the asymptomatic incubation period (7-9). Thus, knowledge of length and dispersion of incubation period is crucial for SARS-CoV-2 prevention and control. In addition, transmission dynamics models are designed to mimic the spread of SARS-CoV-2 in a nonlinear fashion, and are broadly used for long-term forecasting and evaluating the effect of prevention measures (10). However, many parameters associated with SARS-CoV-2 transmission are poorly understood, including the incubation period, resulting in a biased prediction (11). Multiple studies have explored the incubation period for COVID-19, but conclusions remain controversial due to limited sample sizes for each study and considerable heterogeneity between studies (12, 13).

Given the continuing global spread of COVID-19, a further investigation of viral incubation by a systematic review and meta-analysis could provide urgently needed support to improve the understanding of COVID-19 transmission potential and aid prediction and decision-making.

## METHODS

### Data Sources and Searches

In this systematic review and meta-analysis, we searched PubMed, Embase, medRxiv, bioRxiv, and arXiv to identify studies related to COVID-19 published or publicly posted from December 01, 2019 to April 26, 2020 (date of last search) in parallel by two authors (L.W. and Y.L.). Each database was searched using the terms “(COVID-19) OR (2019-nCoV) OR (novel coronavirus pneumonia)”. The search strategy is detailed in the Supplement Table 1–4. There were no language restrictions on the search.

### Study Selection

COVID-19 studies that described either individual-level incubation period data or summarized statistics for central tendency and dispersion measures of incubation period were recruited. Studies were excluded if they met either of the below criteria: (1) irrelevant subject to incubation period; (2) no individual-level incubation period or insufficient summarized statistics for incubation period (central tendency and dispersion measures are required); (3) non-human studies; (4) sample size for incubation analysis less than 5; (5) studies of insufficient quality; (6) ambiguous definition of incubation period. Figure 1 describes the literature searching steps. Duplicate studies and studies irrelevant to incubation period were deleted, and studies identified via reference list searches were added. Two reviewers then selected 10% of the retrieved articles at random and independently reviewed the title and abstract according to the predefined set of exclusion criteria. Exclusion criteria were applied consistently, indicating high concordance. In case of uncertainty about inclusion or exclusion, the reviewers consulted together.

**Figure 1:**
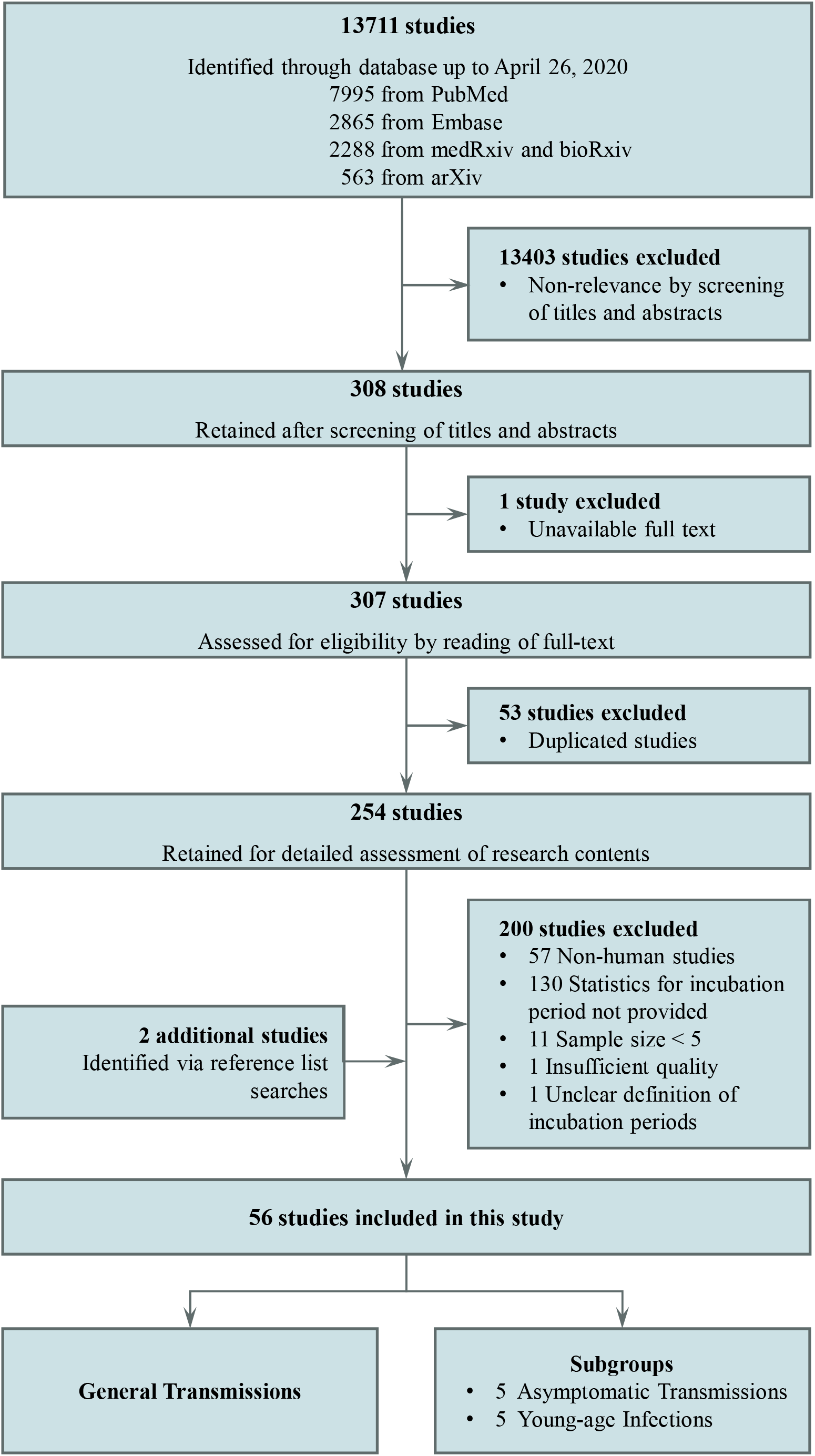
PRISMA flow diagram. PRISMA: Preferred Reporting Items for Systematic Reviews and Meta-Analysis. Fifty-six studies were included in the meta-analysis.

### Data Extraction and Quality Assessment

The literature quality assessment was evaluated in parallel by two researchers (L.W., Y.L.) according to Agency for Healthcare Research and Quality (AHRQ) guidelines (Supplement Table 5). Disagreement between the two researchers was resolved by consensus and the resolution was confirmed by two senior authors (F.C., Y.W.). From each recruited study, either individual-level incubation period data or summarized statistics for central tendency (mean or median) and dispersion (variance, standard deviation, interquatiles, or range) measures were extracted, as well as population characteristics including sample size, average age, and male proportion (Supplement Table 6). Data were extracted by two independent research coordinators from each publication (L.W., Y.L.); inconsistent inputs were verified and justified by a third author to ensure correctness of data extraction (Y.W.). All literature included in the meta-analysis was labeled as “General Transmissions”.

In addition, three publications reported characteristics of incubation among cases infected by asymptomatic or presymptomatic carriers, and two studies had a subset of cases infected by carriers in the asymptomatic period (14, 15); these five studies were labeled as “Asymptomatic Transmissions”. Five studies investigated the incubation period among young-age infections (age < 20 years), which were labeled as “Young-age Infections”.

### Data Synthesis and Analysis

Incubation period was assumed to follow a log-normal distribution (16). Parameters of the log-normal distribution including mean and variation were calculated for each study (Supplement Table 6–12). Funnel plots and Egger tests were used to show the potential publication bias and study heterogeneity. The expected mean of log-scaled incubation period was summarized by meta-analysis followed by exponential calculation to obtain the median of incubation period and the corresponding 95% confidence interval (95%CI). Random-effects meta-analysis was used if *P* value for heterogeneity test ≤ 0.05; otherwise, fixed-effects meta-analysis would be used.

On the other hand, the dispersion for the incubation period was estimated by *e*σ in which the σ is the estimate of standard deviation of the corresponding log-scaled distribution (17). Variances of log-scaled incubation period of the recruited studies were assumed to follow the inverse gamma distribution to estimate the expectation value. Bootstrap was used to estimate the corresponding 95%CI of the dispersion.

Further, the distribution of the incubation period was simulated for the general transmissions, asymptomatic transmissions, and young-age infections, respectively. The 1 000 posterior means of the log-scaled distribution of incubation period were generated using the Bayesian model that produces probability distribution for each parameter, within R package *bayesmeta* (18). In addition, the 1 000 standard deviations of the log-scaled distribution were generated by Bootstrap sampling. The 1 000 means and standard deviations of the log-scaled distribution were used to simulate the distribution of incubation period. The proportion of infections developing symptoms after a certain length of quarantine were estimated; the risk of symptomatic SARS-CoV-2 infections being undetected after a certain length of active monitoring among the active-monitoring population was estimated as well (19, 20).

Last, meta-regression was used to explore the association between the age, sex, and median incubation period. All statistical analyses were performed using R software version 3.6.1 (Foundation for Statistical Computing).

### Role of the Funding Source

This study was funded by the National Natural Science Foundation of China (82041024 to F.C., 82041026 to H.S., 81973142 to Y.W.). Sponsors had no role in design of the study, collection and analysis of data, or preparation of the manuscript.

## RESULTS

Our search retrieved 13 711 records, of which 13 403 were irrelevant to incubation period and were excluded during screening of titles and abstracts (Figure 1). Fifty-three duplicate studies and 1 study that had no access to full text were removed. By careful screening the full text of the remaining 254 studies, we found that 130 with no summarized statistics for incubation period, 57 non-human studies, 11 studies with sample size less than 5, 1 study of insufficient quality, and 1 with ambiguous definition of incubation period were ineligible. Meanwhile, 2 additional studies were identified via reference list searches. Finally, there were 56 studies that met the inclusion criteria, including 34 published studies, 22 preprint studies, and 4 095 COVID-19 infections in total. Literature quality was evaluated for each included study according to AHRQ guidelines (Supplement Table 5). Summarized statistics of incubation periods and population characteristics extracted from each study were shown in Supplement Table 6.

Parameters for log-normal distribution of incubation period were derived for each study (Supplement Figure 1). Visual inspection of the funnel plots showed no risk of publication bias, as confirmed by means of the Egger test (*P* = 0.2877) (Supplement Figure 2). Due to considerable heterogeneity among studies (*I*^2^ = 96.1%, *P* < 0.0001), random-effects meta-analysis was used to estimate the median incubation period to be 5.8 days (95%CI, 5.3–6.2 d) (Figure 2); the corresponding mean incubation period was 6.9 d. Notably, incubation period of general transmissions was shorter than that of asymptomatic transmissions (median, 7.7; 95%CI, 6.3–9.4 d, *P* = 0.0408) and that of young-age infections (median, 7.3 d; 95%CI 6.2–8.6 d, *P* = 0.0219) (Figure 3A, Supplement Figure 3). In addition, summarized results for preprint studies without peer-review (median, 6.4 d; 95%CI, 5.8–7.0 d) showed almost one-day longer of median incubation period than that among studies published in scientific journals (median, 5.4 d; 95%CI, 4.8–6.0 d; *P* = 0.0489) (Figure 3B, Supplement Figure 4). No difference was observed among studies in mainland China versus those performed in regions other than mainland China (*P* = 0.6075) (Figure 3C, Supplement Figure 5). Additionally, rather than solely focusing on the median, the dispersion of the incubation period was studied. Dispersions were estimated as 1.80 (95%CI, 1.59–2.06) for general transmissions, 1.37 (95%CI, 1.24–1.63) for asymptomatic transmissions, and 1.90 (95%CI, 1.55–2.61) for young-age infections, respectively.

**Figure 2:**
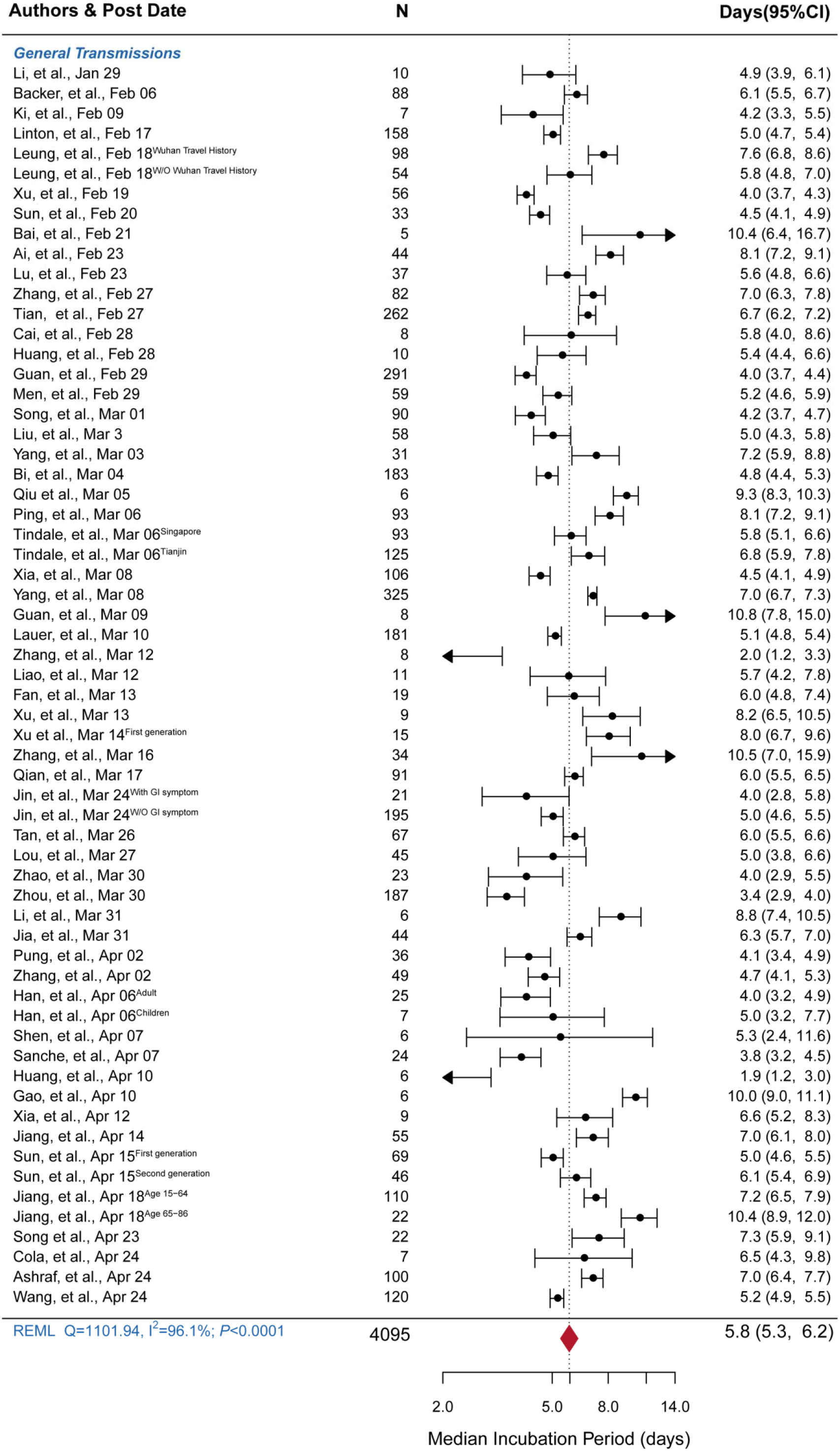
Forest plot for median incubation period among general transmissions. Studies were ordered by date of post online. Significant heterogeneity was observed among studies (*I*^2^ = 96.1%, P < 0.0001). The random-effects meta-analysis using restricted maximum likelihood (REML) was used to summarize the median incubation period (days) and the corresponding 95% confidence interval (95%CI).

**Figure 3:**
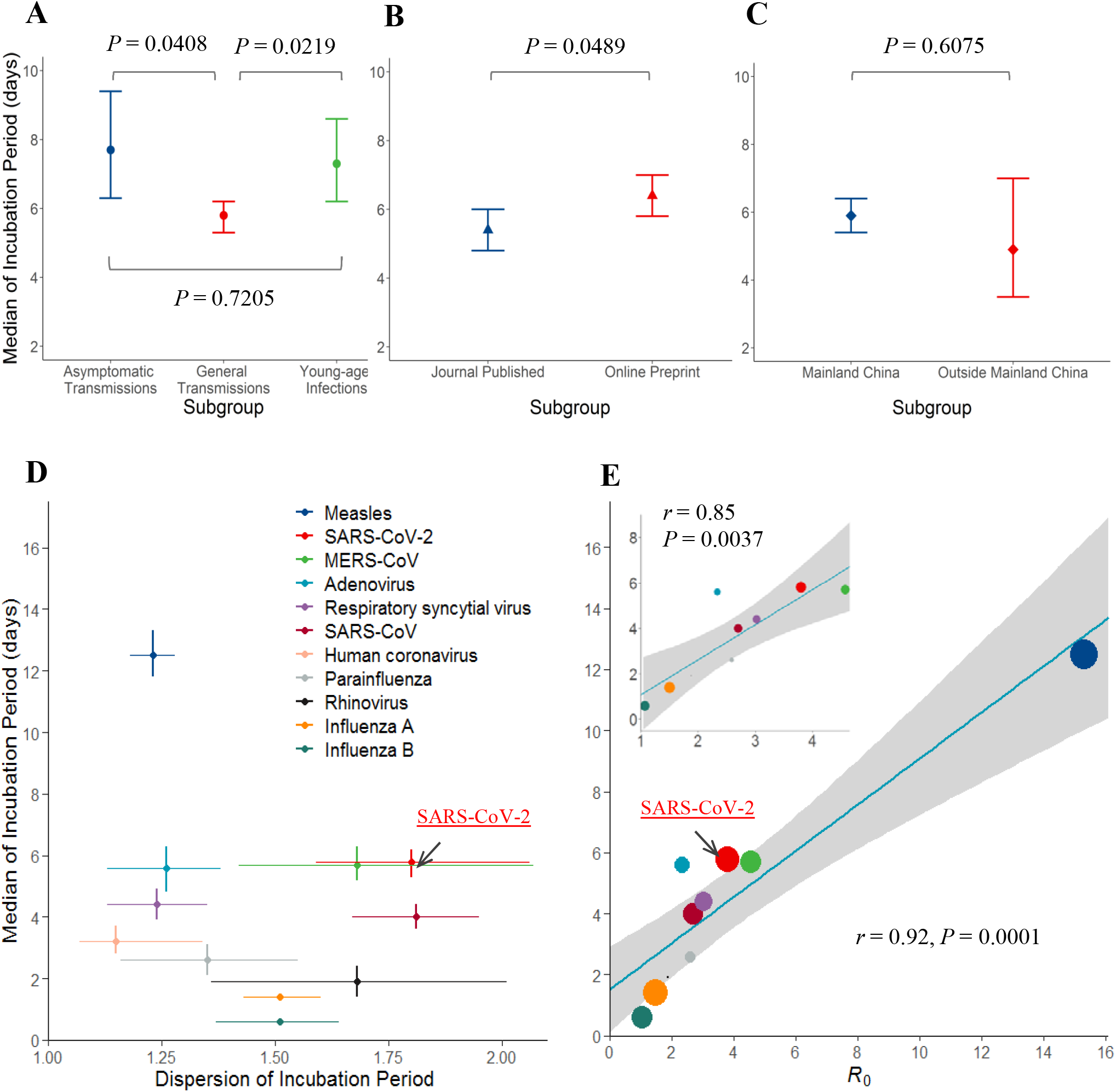
Subgroup meta-analyses, and comparison for characteristics of incubation period among 11 viral respiratory infections. Subgroup meta-analyses were performed among (A): General Transmissions versus Asymptomatic Transmissions versus Young Age Infections, (B): journal-published studies versus preprint studies, and (C): studies performed in mainland China versus those outside mainland China. Median incubation period between the subgroups was compared using Z test in logscale. Median and dispersion of the incubation period, and the corresponding 95% confidence interval for 11 viral respiratory infections are demonstrated (D); black arrow highlights the results for SARS-CoV-2. Correlation between the length of incubation period and basic reproductive number (*R*_0_) was evaluated by linear regression (E); the correlation was reanalyzed by excluding the results of measles in a sensitivity analysis.

For comparison with other viral respiratory infections, the summarized statistics of incubation periods for 9 viral respiratory infections were obtained from a previously published systematic review, including measles, adenovirus, respiratory syncytial virus, SARS-CoV, human coronavirus, parainfluenza, rhinovirus, influenza A, and influenza B (17). In addition, meta-analysis was performed for the incubation period of MERS (Supplement Material). COVID-19 had a significantly longer incubation period than that of SARS (median, 4.0 d; 95% CI, 3.6–4.4 d) (*P* < 0.0001), but similar to that of MERS (median, 5.7 d; 95% CI, 5.2–6.3 d) (*P* = 0.6392) (Figure 3D, Supplement Figure 8). Further, among median incubation periods for 11 viral respiratory infections, SARS-CoV-2 ranked second after measles (Figure 3D). The dispersion of incubation period of SARS-CoV-2 also ranked second (Figure 3D). The basic reproductive numbers (*R*_0_) of the assessed respiratory viruses were strongly correlated with the length of incubation period (*r* = 0.92, *P* = 0.0001) and retained statistical significance by excluding one outlier (*r* = 0.85, *P* = 0.0037) (Figure 3E).

The distribution of incubation period was simulated; 6.7% (95%CI, 2.4–11.2%) and 1.4% (95%CI, 0.1–3.6%) of general transmissions had an incubation period over 14 d and 21 d, respectively (Figure 4A); these proportions reached 14.6% (over 14 d period; 95%CI, 3.7–28.3%) and 4.4% (over 21 d period; 95%CI, 0.0–13.3%) (Figure 4B) among young-age infections due to the relatively large median and dispersion. In addition, 2.9% (95%CI, 0.0–13.0%) and 0.1% (95%CI, 0.0–2.2%) of asymptomatic transmissions had an incubation period over 14 d and 21 d, respectively (Figure 4C). The 97.5^th^ percentiles of incubation period in the population of general transmissions, young age infections, and asymptomatic transmissions were 18 d, 25 d, and 14 d, respectively (Figure 4D, 4E, 4F).

**Figure 4:**
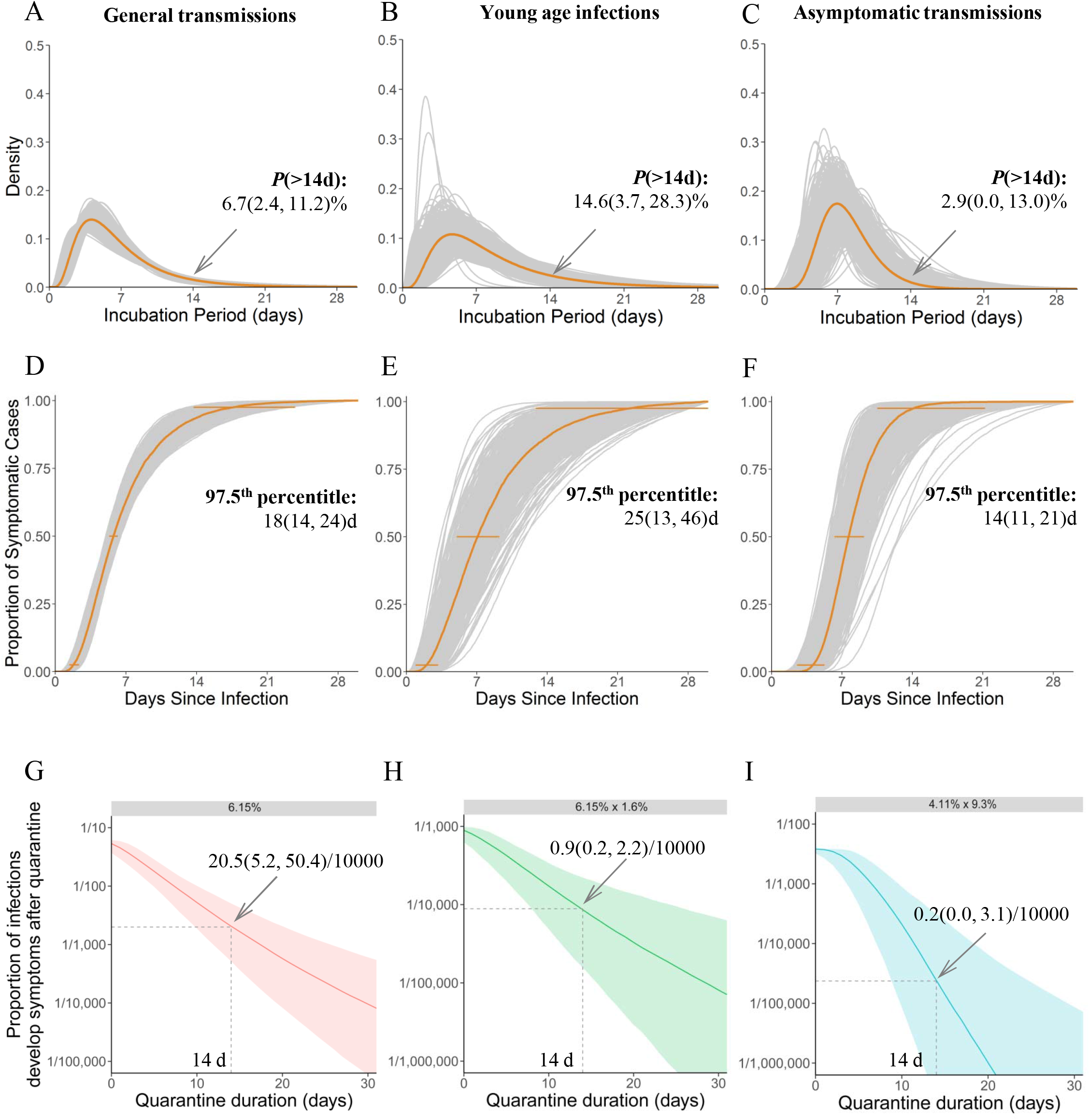
Distributions of incubation period and risk evaluation for the length of quarantine period. Density distributions of the incubation period for general transmissions (A), young age infections (B), and asymptomatic transmissions (C) are shown, respectively; the proportion and the 95% confidence interval (95%CI) of infections having incubation period over 14 days were estimated. Cumulative distributions of incubation period for general transmissions (D), young-age infections (E), and asymptomatic transmissions (F) are shown, respectively; the 97.5^th^ percentile and the 95%CI were estimated for each distribution. In addition, assuming a 6.15% risk of being infected among the general quarantine population, the risk of having infections develop symptoms after 14-day active monitoring or quarantine was estimated (G). Further, assuming the young age infections account for 1.6% of general transmissions, the probability of having young age infections among the general quarantine population was set at 6.15%×1.6%; the risk of having infections develop symptoms after 14-day quarantine period among the quarantine population was estimated (H). Similarly, assuming that individuals who contacted latent infections during the asymptomatic or presymptomatic period, which account for 9.3% of the general quarantine population, and 4.11% probability to be infected, the risk of observed asymptomatic transmissions developing symptoms after 14-day quarantine period is shown in Panel (I).

According to the results of an epidemiological study of an active monitoring population in China (Supplement Table 14) (21), assuming a 6.15% risk of symptomatic infections among the overall active monitoring population with close contacts, the estimated probability of symptomatic SARS-CoV-2 infections that would be undetected after 14-day active monitoring or quarantine was 20.5 (95%CI,5.2–50.4) per 10 000 monitored individuals (Figure 4G). Similarly, one study summarized the proportion of young-age infections at 1.6% among general infections (22); thus, among the overall active monitoring population, the estimated probability of symptomatic SARS-CoV-2 infections that would be undetected after 14-day active monitoring was 0.9 in 10 000 (95%CI: 0.2–2.2 in 10 000) (Figure 4H). In addition, assuming a 4.11% risk of symptomatic infections among contacts with latent infections and a proportion of latent infections at 9.3% in the overall active monitoring population, the estimated probability of undetected SARS-CoV-2 infections with no sympotom within 14-day active monitoring was 0.2 in 10 000 (95%CI: 0.0–3.1 in 10 000) (Figure 4I). Sensitivity analyses considering various settings for the risk of being infected among an active monitoring population were performed as well (Supplement Table 14). Overall, in the active monitoring population, the risk of developing symptoms after a 14-day quarantine period was about 20 in 10 000, while the risk of young age infections and asymptomatic transmissions developing symptoms after 14-day quarantine period was below 1 in 10 000.

Average age and male proportion were extracted from 24 of 56 studies. Meta-regression incorporating two moderators simultaneously was used to explore the impact of individual characteristics on length of incubation period. A linear relationship was identified between age and log-scaled median of incubation period. Average age per 10-year increments resulted in a 16% increment in median incubation period with adjustment for male proportion (incubation period ratio, 1.16, 95%CI 1.01–1.32; *P* = 0.0250) (Supplement Figure 6A). No evidence indicated an association between sex and median of incubation period (*P* = 0.1315) (Supplement Figure 6B).

Finally, an interactive real-time risk assessment application was developed to provide real-time updates of the risk assessment for symptomatic SARS-CoV-2 infections that would be undetected during active monitoring among active monitoring population with close contacts by setting several crucial parameters (Supplement Figure 10).

## DISCUSSION

Increasing evidence supports the transmission potential of SARS-CoV-2 during the latent period (7-9). Thus, length of incubation period is a crucial parameter to determine the risks for close contacts and guide contact tracing and quarantine policies. The estimated median incubation period in this study was 5.8 days for general transmissions; the estimated mean incubation was 6.9 days which is 33% longer than the previously frequently adopted value—5.2 mean days as reported by Li (16). Notably, asymptomatic transmissions and young-age infections appear to have an almost two-day longer incubation period than general transmissions. Infections contacted with latent infections may have a low viral load that requires a longer incubation to develop symptoms (23, 24). On the other hand, young-age infections probably have a strong immune status that results in a longer incubation. Interestingly, preprint studies appear to report longer incubation period than do journal publications, indicating potential publication bias. However, large variation of estimates was observed among studies, which indicates a non-negligible heterogeneity in COVID-19 patients. Evolution of SARS-CoV-2 might partially address this heterogeneity (25).

Our study demonstrates that SARS-CoV-2 has a considerably longer incubation period than most types of viral respiratory infections. Notably, a significant positive association between length of incubation period and magnitude of *R*_0_ was observed. This finding indicates viral respiratory infections beyond SARS-CoV-2 may have transmission potential during their incubation period. In addition, SARS-CoV-2 has a high dispersion of incubation period, which increases the difficulty in tracing and controlling for contacts. These unique epidemiologic characteristics partially contribute to today’s global spread of COVID-19.

The 14-day quarantine period has been adopted in mainland China and suggested to the international community by WHO (26). We estimated that 7 out of 100 general infections would develop symptoms after 14 days. Further, nearly 15 out of 100 infected people under age 20 years will develop symptoms after 14 days. However, considering the probability of being infected among contacts of the population in active monitoring or quarantine, about 20 per 10 000 contacts would develop symptoms after 14 days in active monitoring or quarantine; the risk of observing young-age infections or asymptomatic transmissions developing symptoms after 14 days is below 1 in 10 000. Assuming the risks of being infected for the active monitoring population having contacts with symptomatic infections and latent infections were 1% in equal, about 3 in 10 000 contacts would develop symptoms after 14 days, which is more than 1.0 in 10 000 reported in the previous study.^19^ Thus, the 14-day quarantine policy is sufficient to trace and identify infections among an active monitoring population. However, precise understanding of the crucial epidemiological parameters related to transmission probability in active monitoring population could aid in further refining the appropriate length of quarantine (27).

In addition, age is likely to have a positive relationship with the length of incubation period. Notably, the result in our study indicated that older adults had a longer incubation period than younger adults, which was consistent with the findings of previous studies (28, 29). Older adults tend to have more health complications such as respiratory issues and chronic diseases; thus, pre-existing symptoms may mask the onset of COVID-19 symptoms, which could bias the measurement of incubation period. However, the underlying mechanism is unclear and warrants further investigation.

We acknowledge some limitations of this study. First, the sample sizes for asymptomatic transmissions and young-age infections are small, and the results for these subgroups may be less representative. Second, most studies obtained data from public resources, and the raw data were not provided; there is a possibility that some data were used repeatedly. Third, 50 of 56 studies were from mainland China; studies from other regions and countries are needed to explore the impact of viral evolution on variation of incubation period and other epidemiological characteristics. Fourth, precisely estimating the exposure window and time of symptom onset related to SARS-CoV-2 infection could be difficult in practice. Studies used different methods to quantify the uncertainty of incubation period for each individual, which may partially explain the non-negligible intra-study heterogeneity. Last, knowledge of the risk of being infected among close contacts is limited and may vary due to different definition of close contacts.

## CONCLUSIONS

In conclusion, this study integrated 56 studies and 4 095 COVID-19 infections and estimated the median and dispersion of the SARS-CoV-2 incubation period, both of which ranked second among 11 viral respiratory infections. A long and dispersive incubation period probably contributes to the increasing spread of COVID-19 worldwide. Yet, the 14-day quarantine period is sufficient to trace and identify symptomatic infections among an active monitoring population, and monitoring and quarantine policies may be adjusted to accommodate higher-risk age groups.

## Data Availability

Additional data available from the corresponding author at upon reasonable request.

## Author Contributors

Y. Wei and F. Chen had full access to all of the data in the study and take responsibility for the integrity of the data and the accuracy of the data analysis.

Concept and design: Y. Wei, F. Chen, H. Shen.

Acquisition, analysis, or interpretation of data: L. Wei, Y. Liu, J. Chen, Y. Zhao, L. Huang, Y. Wei.

Drafting of the manuscript: Y. Wei, L. Wei.

Critical revision of the manuscript for important intellectual content: H. Shen, F. Chen, Y. Wei.

Statistical analysis: L. Wei, Y. Liu, R. Zhang, Y. Zhao, J. Chen, L. Huang, Y. Wei.

Obtained funding: Y. Wei, F. Chen, H. Shen.

Administrative, technical, or material support: F. Chen, H. Shen. Supervision: Y. Wei, F. Chen, H. Shen.

Approval of final report: All authors.

## Funding

This study was funded by the National Natural Science Foundation of China (82041024 to F.C., 82041026 to H.S., 81973142 to Y.W., 81903407 to L.H.). Sponsors had no role in design of the study, collection and analysis of data, or preparation of the manuscript.

## Conflicts of Interest

The authors have declared that no conflict of interest exists. All authors have completed the ICMJE uniform disclosure form at www.icmje.org/coi_disclosure.pdf and declare: no support from any organization for the submitted work; no financial relationships with any organizations that might have an interest in the submitted work in the previous three years; no other relationships or activities that could appear to have influenced the submitted work.

## Notes

### Competing Interest Statement

The authors have declared no competing interest.

